# Impact of Code Stroke Activation and Warfarin Use on Time to Anticoagulation Reversal in Intracerebral Hemorrhage: Implications For Quality Improvement

**DOI:** 10.64898/2026.07.29.26359285

**Authors:** Heidi Tan, Michael Carrillo, Diana Dench, Jefferson Chen, Mohammad Shafie, Wengui Yu

**Affiliations:** Department of Neurology, University of California, Irvine; Department of Neurosurgery, University of California, Irvine

**Keywords:** Intracerebral hemorrhage, warfarin, direct oral anticoagulants, anticoagulation reversal, door-to-treatment time, Code Stroke, 4 factor-prothrombin complex concentrates

## Abstract

**Background:** Spontaneous intracerebral hemorrhage (ICH) is the most devastating type of stroke but lacks the established time-critical treatment guidelines available for acute ischemic stroke. This study investigated the impact of Code Stroke activation and warfarin use on anticoagulation reversal times to identify opportunities for quality improvement.

**Methods:** We retrospectively assessed patients with anticoagulation-associated ICH admitted to our medical center between January 1, 2020, and December 31, 2025. Patients with warfarin- or direct oral anticoagulant (DOAC)-associated ICH were identified from our ICH clinical trial screening log, Vizient, and AHA’s *Get With The Guidelines-Stroke* registry. Code Stroke activation, warfarin or DOAC use, anticoagulation reversal times, causes of delay, and clinical outcomes at hospital discharge were analyzed.

**Results:** Of 892 ICH admissions, 50 patients had confirmed anticoagulation-associated ICH. Among those who underwent reversed at our center (n=34), Code Stroke activation (n=24) significantly reduced door-to-CT time (16.5 [12.8-22.0] vs 172.5 [49.0-261.5] minutes, p <0.001), reversal agent order-to-needle time (37.5 [24.7-56.9] vs 57.0 [39.0-85.0] minutes, p <0.001), and door-to-treatment (DTT) time (64 [48– 98] vs 277 [159–305] minutes, *p* <0.001) compared to non-activation (n=10). Conversely, warfarin use was associated with higher international normalized ratios (INR) (2.7 [2.2-3.6] vs 1.3 [1.2-1.9], *p* =0.003) and significantly longer DTT time (95 [51-108] vs 64 [48-77] minutes, *p* =0.007). Primary DTT delays stemmed from the absence of a time-critical treatment protocol, weight-based dosing for 4F-PCC, waiting for INR results, and a lack of Code Stroke activation for patients with mild symptoms, trauma or unexplained unresponsiveness.

**Conclusions:** Our findings suggest that Code Stroke activation for all suspected cases of ICH, a time-critical emergency department algorithm targeting a DTT time of less than 60 minutes, and immediate anticoagulation reversal with fixed-dose 4F-PCC without waiting for INR results may optimize the acute management of anticoagulation-associated ICH.

## INTRODUCTION

Spontaneous intracerebral hemorrhage (ICH) accounts for 28.8% of all strokes worldwide.^[1]^ Uncontrolled hypertension, cerebral amyloid angiopathy, and oral anticoagulant use are the most common causes of ICH.^[2–6]^ Hematoma expansion is an independent predictor of poor functional outcome.^[7–8]^ Several randomized controlled trials (RCTs) of intensive blood pressure control, emergent anticoagulation reversal, or hemostatic therapy with recombinant activated factor VII (rFVIIa) have demonstrated reductions in hematoma expansion without significant improvement in functional outcomes.^[8–14]^ However, a few recent studies have showed that ultra-early bundled care, including simultaneous blood pressure control, anticoagulation reversal within 60 minutes, or minimal hematoma evacuation, reduces the risk of hematoma expansion and improves functional outcome.^[15–18]^

Anticoagulation-associated ICH is linked to a higher risk of hematoma expansion and mortality.^[4,5]^ Among all stroke subtypes, warfarin-associated ICH carries the highest mortality (43-52%).^[4,6]^ Despite these devastating outcomes, ICH lacks established, time-critical management guidelines in contrast to acute ischemic stroke (AIS).[^19–21]^ This consensus view prompted experts in the field to call for a standardized “Code ICH” protocol.^[21]^

In a retrospective study using data from 11 comprehensive stroke centers participating in the American Heart Association’s *Get With The Guidelines (GWTG) Stroke* registry, the time-to-treatment for patients with ICH was significantly longer than for patients with AIS.^[22]^ Only 37% of patients with anticoagulation-associated ICH received anticoagulation reversal with a door-to-treatment (DTT) time ≤ 90 minutes. Similarly, in a cohort study of 7469 patients with anticoagulation-associated ICH from 465 US hospitals participating in the *GWTG*-*Stroke* registry, only 27.7% patients achieved a DTT time ≤ 60 minutes.^[23]^ Notably, a DTT time of ≤ 60 minutes for anticoagulation reversal was associated with decreased mortality and discharge to hospice. These findings underscore the critical need to identify factors contributing to DTT delays and to optimize the acute management of anticoagulation-associated ICH.

We implemented a Code Stroke algorithm incorporating anticoagulation reversal in 2015.^[24,25]^ This study aimed to evaluate the impact of Code Stroke activation and warfarin use on DTT times for anticoagulation reversal following ICH and to identify concrete opportunities for quality improvement.

## METHODS

### Study Design and Ethical Approval

This retrospective study was approved by the Institutional Review Board (IRB) of the University of California, Irvine. The requirement for informed consent was waived due to the retrospective study design and the minimal risk posed to participants. All methods were conducted in accordance with applicable regulations and the STROBE reporting guidelines for observational studies. De-identified data are available from the corresponding author upon reasonable request.

### Patient population

Consecutive patients with spontaneous ICH admitted to the University of California, Irvine Medical Center between January 1, 2020, and December 31, 2025, were screened for inclusion. The patient list, including demographic information and admission dates, was generated from our program’s screening log for the Anticoagulation in Intracerebral Hemorrhage Survivors for Stroke Prevention and Recovery (ASPIRE) trial, the Vizient database, and the AHA’s *GWTG*-*Stroke* registry.^[26–27]^ All patients with suspected warfarin- or direct oral anticoagulant (DOAC)-associated ICH were included for manual chart review by 2 co-authors (H.T. and M.C.) and independent verification by senior author (W.Y.).

The following data were abstracted from the electronic medical record (EPIC): age, race, past medical history, home medications, indication for anticoagulation, time of last anticoagulant dose, time of symptom onset or last-known-well, emergency department (ED) arrival time, National Institutes of Health Stroke Scale (NIHSS) score, ICH score, international normalized ratio (INR), onset-to-treatment time (OTT), DTT time for anticoagulation reversal, and functional outcome at hospital discharge as assessed by the modified Rankin Scale (mRS).

All patients underwent standard diagnostic evaluation and treatment, including anticoagulation reversal in accordance with treatment guidelines.^[24,25,28]^ Our institutional protocol for anticoagulation reversal includes intravenous (IV) four-factor prothrombin complex concentrates (4F-PCC; 25–50 units/kg) for factor Xa inhibitor–associated ICH, idarucizumab for dabigatran-associated ICH, and IV 4F-PCC (25–50 units/kg) combined with IV vitamin K (10 mg) for warfarin-associated ICH, with dosing guided by the INR.^[25]^ An additional dose of 4F-PCC and/or vitamin K was administered if the repeat INR remained elevated.

### Statistical analysis

Continuous variables with a normal distribution were described by mean ± standard deviation (SD). Non-normally distributed variables were reported as median with interquartile ranges (IQR). Categorical variables were expressed by counts and percentages. Baseline characteristics and functional outcomes at hospital discharge were compared between study groups using the independent student T test or the Mann-Whitney U test as appropriate. Due to the small sample size, Fisher’s exact test was used to evaluate categorized data. Statistical significance was set at p < 0.05. Multivariable logistic regression modeling was not performed due to sample size constraints. All statistical analyses were conducted using SPSS version 29.0.

## RESULTS

Among 892 consecutive patients admitted with spontaneous ICH to our medical center between January 1, 2020, and December 31, 2025, 103 were initially flagged as having suspected anticoagulation-associated ICH. Fifty patients (5.6%) were confirmed to have true anticoagulation-associated ICH via manual chart review and independent verification (Figure 1). Fifty-three patients were excluded for the following reasons: not actively taking anticoagulants (n=35), traumatic intracranial hemorrhage (n=9), hemorrhagic transformation of an ischemic stroke (n=5), brain metastasis (n=1), severe thrombocytopenia (n=1), cerebral venous sinus thrombosis (n=1), or hemorrhagic stroke from infectious endocarditis (n=1).

**Fig 1.**
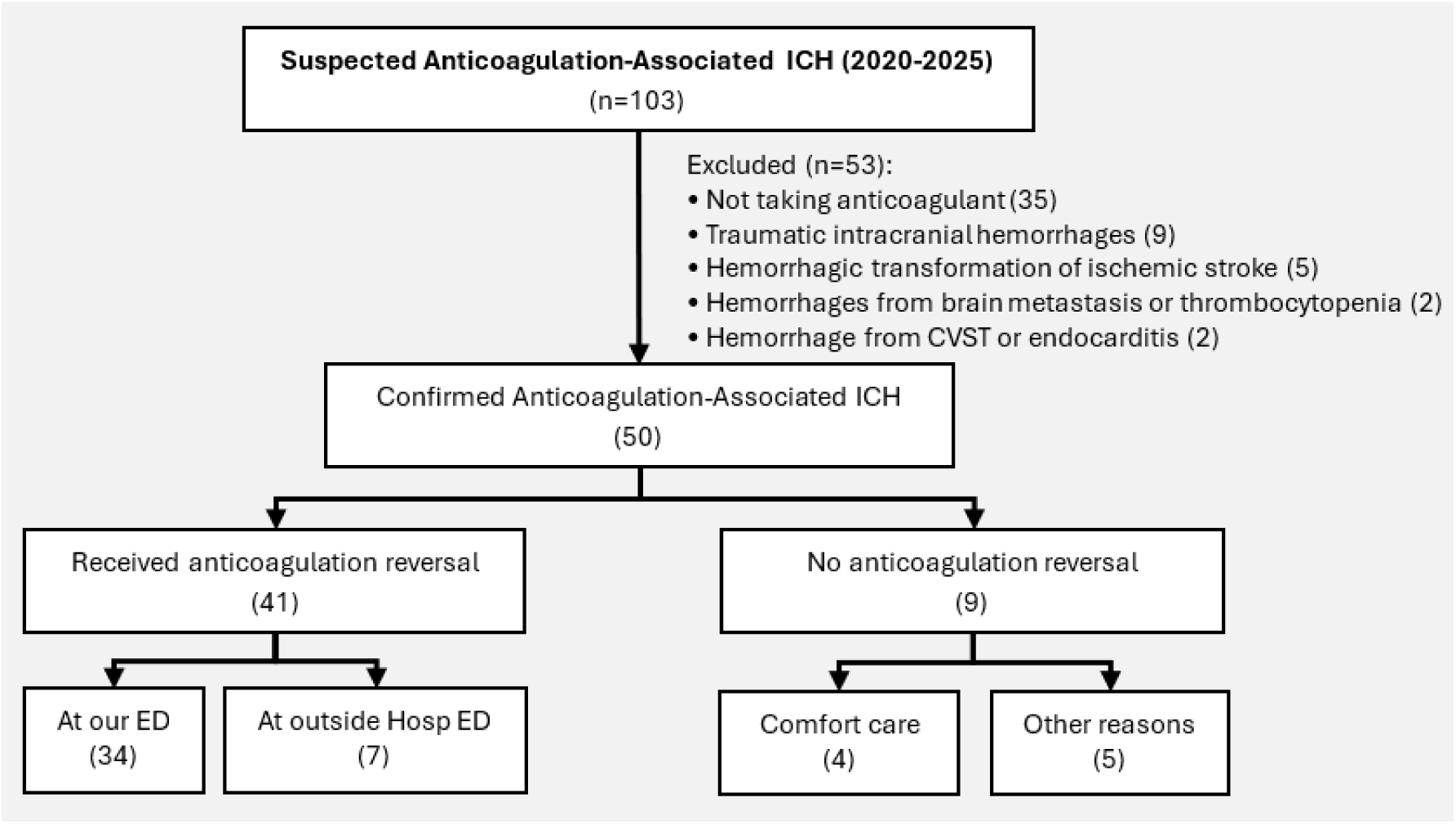
The study flowchart

Among the 50 patients with confirmed anticoagulation-associated ICH, the most common comorbidities were hypertension (92%), atrial fibrillation or atrial flutter (78%), and hyperlipidemia or dyslipidemia (52%). Apixaban was the most frequently used anticoagulant (60%), followed by warfarin (22%) and rivaroxaban (16%). Forty-one patients (82%) received anticoagulation reversal therapy, whereas nine patients (18%) did not.

As shown in Table 1, there were no statistically significant differences between the reversal and non-reversal groups regarding demographics, past medical history, type of oral anticoagulant, NIHSS score, ICH score, INR, or functional outcomes at hospital discharge. Twenty-seven patients received 4F-PCC alone for anticoagulation reversal, 10 received 4F-PCC plus vitamin K, 2 received 4F-PCC plus fresh frozen plasma (FFP), 2 received low-dose vitamin K alone for warfarin-associated ICH with an INR < 2.0. One patient with a history of atrial fibrillation on Eliquis and end-stage-renal disease (ESRD) on hemodialysis developed an acute left arm arteriovenous (AV) fistula clot and an occlusive deep vein thrombosis (DVT) in the left brachiocephalic and subclavian veins following anticoagulation reversal with 4F-PCC at an outside facility. An additional 3 patients developed nonocclusive, indwelling catheter-associated DVTs in either the common femoral vein (n=2) or internal jugular vein (n=1). No cases of pulmonary embolism, acute ischemic stroke, or myocardial infarctions occurred in patients who received anticoagulation reversal.

**Table 1.**
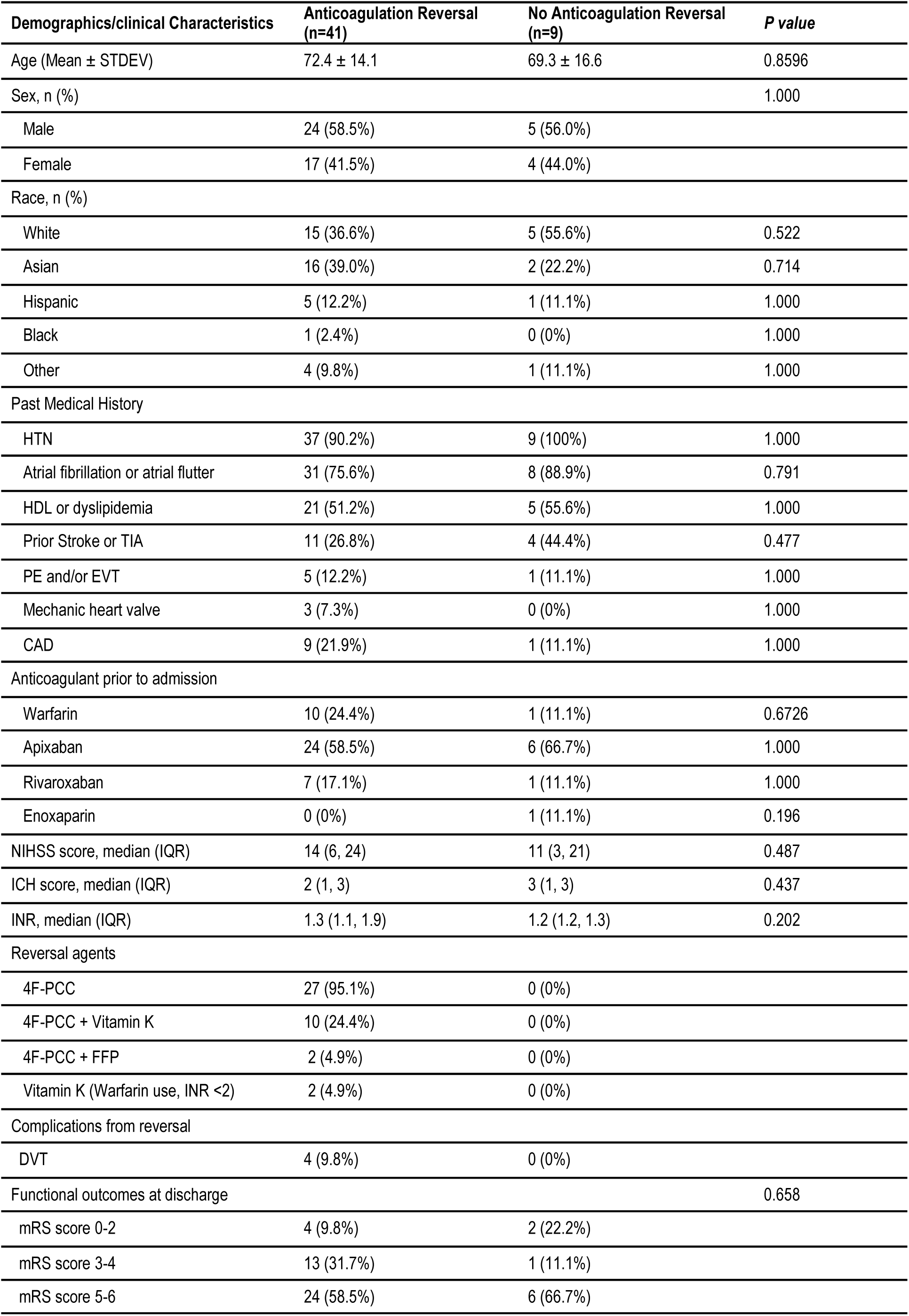
Clinical characteristics of anticoagulation-associated ICH with or without anticoagulation reversal.

| Demographics/clinical Characteristics | Anticoagulation Reversal (n=41) | No Anticoagulation Reversal (n=9) | *P* value |
| --- | --- | --- | --- |
| Age (Mean ± STDEV) | 72.4 ± 14.1 | 69.3 ± 16.6 | 0.8596 |
| Sex, n (%) |  |  | 1.000 |
| Male | 24 (58.5%) | 5 (56.0%) |  |
| Female | 17 (41.5%) | 4 (44.0%) |  |
| Race, n (%) |  |  |  |
| White | 15 (36.6%) | 5 (55.6%) | 0.522 |
| Asian | 16 (39.0%) | 2 (22.2%) | 0.714 |
| Hispanic | 5 (12.2%) | 1 (11.1%) | 1.000 |
| Black | 1 (2.4%) | 0 (0%) | 1.000 |
| Other | 4 (9.8%) | 1 (11.1%) | 1.000 |
| Past Medical History |  |  |  |
| HTN | 37 (90.2%) | 9 (100%) | 1.000 |
| Atrial fibrillation or atrial flutter | 31 (75.6%) | 8 (88.9%) | 0.791 |
| HDL or dyslipidemia | 21 (51.2%) | 5 (55.6%) | 1.000 |
| Prior Stroke or TIA | 11 (26.8%) | 4 (44.4%) | 0.477 |
| PE and/or EVT | 5 (12.2%) | 1 (11.1%) | 1.000 |
| Mechanic heart valve | 3 (7.3%) | 0 (0%) | 1.000 |
| CAD | 9 (21.9%) | 1 (11.1%) | 1.000 |
| Anticoagulant prior to admission |  |  |  |
| Warfarin | 10 (24.4%) | 1 (11.1%) | 0.6726 |
| Apixaban | 24 (58.5%) | 6 (66.7%) | 1.000 |
| Rivaroxaban | 7 (17.1%) | 1 (11.1%) | 1.000 |
| Enoxaparin | 0 (0%) | 1 (11.1%) | 0.196 |
| NIHSS score, median (IQR) | 14 (6, 24) | 11 (3, 21) | 0.487 |
| ICH score, median (IQR) | 2 (1, 3) | 3 (1, 3) | 0.437 |
| INR, median (IQR) | 1.3 (1.1, 1.9) | 1.2 (1.2, 1.3) | 0.202 |
| Reversal agents |  |  |  |
| 4F-PCC | 27 (95.1%) | 0 (0%) |  |
| 4F-PCC + Vitamin K | 10 (24.4%) | 0 (0%) |  |
| 4F-PCC + FFP | 2 (4.9%) | 0 (0%) |  |
| Vitamin K (Warfarin use, INR <2) | 2 (4.9%) | 0 (0%) |  |
| Complications from reversal |  |  |  |
| DVT | 4 (9.8%) | 0 (0%) |  |
| Functional outcomes at discharge |  |  | 0.658 |
| mRS score 0-2 | 4 (9.8%) | 2 (22.2%) |  |
| mRS score 3-4 | 13 (31.7%) | 1 (11.1%) |  |
| mRS score 5-6 | 24 (58.5%) | 6 (66.7%) |  |

Of the 9 patients who did not receive anticoagulation reversal, 4 were transitioned to comfort or palliative care following family discussion in the ED. These patients presented with severe ICH accompanied by brain herniation and hydrocephalus, alongside advanced age and/or pre-existing dementia. The remaining 5 patients did not receive anticoagulation reversal because of small hemorrhage volume or because their last anticoagulant dose fell outside the recommended therapeutic reversal window (Table 2).

**Table 2.** The reasons anticoagulation reversal was not administered.

| Reasons | n |
| --- | --- |
| Comfort care or palliative care in ED | 4 |
| Stable neurological examination from small hemorrhages | 2 |
| Last anticoagulant dose outside recommended reversal window | 3 |
| Apixaban > 24 hours since last dose | (2) |
| LMWH > 12 hours since last dose | (1) |

Among the 41 patients who received anticoagulation reversal, 7 were treated at an outside hospital ED before transfer to our medical center for a higher level of care. Of the remaining 34 patients who presented directly to our ED, 24 (70.6%) had a Code Stroke activation upon arrival, whereas 10 did not. Reasons for the lack of Code Stroke activation included mild symptoms (n=5), trauma (e.g., a ground-level fall, being found down, or unresponsiveness on arrival, n=4), and symptom onset exceeding 24 hours (n=1). The Non-Code Stroke group was significantly younger than the Code Stroke group (65.0 ± 14.5 vs 75.9 ± 11.8, *p* = 0.028), likely due to a few young patients presenting with mild symptoms (Table 3).

**Table 3.** Times to anticoagulation reversal between Code Stroke activation and No Code Stroke Activation groups.

|  | Code Stroke Activation<br>(n=24) | No Code Stroke Activation<br>(n=10) | *P* value |
| --- | --- | --- | --- |
| Age, years | 75.9 ± 11.8 | 65.0 ± 14.5 | 0.028 |
| NIHSS score (IQR) | 16.0 (8.8-23.3) | 9.5 (1.3-24.0) | 0.316 |
| ICH score (IQR) | 2 (1-3) | 1.5 (1-4) | 0.653 |
| Indication for AC |  |  |  |
| A Fib/A Flutter | 19 (79.2%) | 8 (80%) | 1.000 |
| AVR, PE or DVT | 5 (20.8%) | 2 (20%) | 1.000 |
| Warfarin Use | 7 (29.2%) | 2 (20%) | 1.000 |
| DOAC Use | 17 (70.8%) | 8 (80%) | 0.547 |
| INR (IQR) | 1.69 (1.2-2.2) | 1.21 (1.2-1.4) | 0.108 |
| Door to CT time (IQR), min | 16.5 (12.8-22.0) | 172.5 (49.0-261.5) | < 0.001 |
| Door-to-needle time (IQ, min | 37.5 (24.7-56.9) | 57.0 (39.0-85.0) |  |
| OTT (IQR), min | 160 (113-316) | 734 (652-1216) | < 0.001 |
| DTT (IQR), min | 64 (48-98) | 277 (159-305) | <0.001 |
| mRS score at discharge | 5 (4-6) | 5 (4-6) | 0.798 |

**Abbreviations:** AC: anticoagulation; AVR: aortic valve replacement; DOAC: direct oral anticoagulant; DTT: door-to-treatment time; DVT: deep vein thrombosis, INR: international normalized ratio; IQR: interquartile ranges; PE: pulmonary embolism; OTT: onset-to-treatment time.

There were no significant differences in clinical severities (measured by NIHSS and ICH scores), indications for anticoagulation, anticoagulant type (warfarin vs DOAC), INR, or mRS scores at hospital discharge between the Code Stroke activation and non-activation groups. However, patients with Code Stroke activation on arrival had statistically significantly shorter door-to-CT times (16.5 [12.8-22.0] vs. 172.5 [49.0-261.5] minutes, p <0.001), reversal agent order-to-needle times (37.5 [24.7-56.9] vs. 57.0 [39.0-85.0] minutes, p <0.001), OTT time (160 [113–316] vs. 734 [652–1216] minutes, p < 0.001), and DTT time (64 [48–98] vs. 277 [159–305] minutes, p < 0.001) compared to those without Code Stroke activation. The shorter DTT time in the Code Stroke group was mostly attributable to reduction in both door-to-CT and reversal agent order-to-needle times. Notably, the median reversal agent order-to-needle time was 37.5 minutes, likely due to the delay from the mixing and preparing weight-based dosing for 4F-PCC.^[29]^

In the Code Stroke activation group, 9 patients (37.5%) achieved a DTT time of less than 60 minutes. Anticoagulation reversal was ordered by ED physicians (n=11, 45.8%), Neurology/Stroke residents (n=7, 29.2%), and Neurosurgery residents (n=6, 25.0%), highlighting the need for a standardized, time-critical ICH management protocol across the multidisciplinary Code Stroke team.

Table 4 compares reversal times between the Warfarin and DOAC groups. There were no significant differences in patient ages, NIHSS scores, ICH scores, or median onset-to-door time between the 2 groups. However, the warfarin group had significantly higher INR values (2.7 [2.2–3.6] vs. 1.3 [1.2–1.9], p =0.003) and longer median DTT times for anticoagulation reversal (95 [51–108] vs 64 [48–77] minutes, p < 0.007) than the DOAC group. This difference was primarily driven by logistical delays associated with waiting for current laboratory INR results before preparing and administering variable, weight-based doses of 4F-PCC and/or vitamin K.

**Table 4.** Times to anticoagulation reversal between Warfarin and DOAC groups.

|  | Warfarin group<br>(n=7) | DOAC Groups<br>(n=17) | *p-value* |
| --- | --- | --- | --- |
| Age, years | 71.0 ± 12.6 | 79.9 ± 11.2 | 0.214 |
| Median NIHSS score (IQR) | 18 (9.5-23.5) | 22 (9.0-23.0) | 0.923 |
| Median ICH score (IQR) | 1 (0-2) | 3 (1-3) | 0.096 |
| INR (IQR) | 2.7 (2.2-3.6) | 1.3 (1.2-1.9) | 0.003 |
| Median OTT (IQR), minutes | 140 (123-449) | 165 (114-302) | 0.949 |
| Median DTT (IQR), minutes | 95 (51-108) | 64 (48-77) | 0.007 |

**Abbreviations:** DOAC: direct oral anticoagulant; DTT: door-to-treatment time; INR: international normalized ratio; IQR: interquartile ranges; PE: pulmonary embolism; OTT: onset-to-treatment time.

## DISCUSSION

Among the 892 ICH admissions at our comprehensive stroke center between January 1, 2020, and December 31, 2025, 50 (5.6%) were confirmed cases of anticoagulation-associated ICH, and 41 (4.6%) presented directly to our facility. This direct-arrival rate for anticoagulation-associated ICH was slightly higher than the estimated rate of 3.6% observed across the entire *GWTG Stroke* registry database between 2015 and 2021.^[23]^

In this single-center study, we identified several potential opportunities for quality improvement in the current management of anticoagulation-associated ICH. Although Code Stroke activation was associated with significantly shorter median DTT time for anticoagulation reversal, treatment remained substantially delayed compared to historical benchmarks for IV thrombolysis in AIS, particularly among patients with warfarin-associated ICH. The primary drivers of delayed anticoagulation reversal included: the absence of a time-critical treatment protocol specific for ICH, a failure to activate Code Stroke on arrival in a substantial proportion of eligible patients (29.4%), the prolonged preparation time required for weight-based 4F-PCC dosing and waiting for laboratory INR results in patients with warfarin-associated ICH.

While our single-center study was underpowered to detect differences in discharge clinical outcomes, a large cohort study of 7469 patients with anticoagulation-associated ICH from 465 US hospitals participating in the *GWTG*-*Stroke* registry demonstrated that rapid anticoagulation reversal (DTT ≤ 60 minutes) was independently associated with reduced mortality and lower rates of discharge to hospice.^[23]^ These findings underscore the urgent need to develop a time-critical treatment protocol for ICH with the same programmatic urgency commonly implemented for patients with AIS.

Although Andexanet alfa provides superior hemostasis in patients with factor Xa inhibitor-associated ICH, it carries a significantly higher risk of thromboembolic complications without establishing improved functional outcomes compared to 4F-PCC.^[11,30]^ Conversely, data from *GWTG-Stroke* showed that the reversal of subtherapeutic warfarin in patients presenting with acute spontaneous ICH and a baseline INR of 1.5 to 1.9 did not improve functional outcomes.^[31]^ Together, these findings suggest that rapid, rather than completely normalized, correction of coagulation parameters is the critical factor in managing anticoagulation-associated ICH.

In a study of 46 patients with intracranial hemorrhage, a fixed-dose 4F-PCC regimen (1,500 units) combined with 10 mg IV vitamin K achieved more rapid warfarin reversal without increasing hospital length of stay, mortality, or redosing requirements compared to a weight-based regimen.^[32]^ The time required to prepare and mix weight-based 4F-PCC dosing can range from 20 to 45 min depending on site-specific pharmacy or ED logistics.^[29]^ Our study showed that the median reversal agent order-to-needle time was 37.5 [24.7-56.9] minutes in the Code Stroke group. This delay could be reduced significantly if institutional protocols transitioned from weight-based 4F-PCC dosing to fixed-dose strategies for anticoagulation reversal. Multiple studies spanning patients with major intracranial and gastrointestinal bleeding have similarly demonstrated that a fixed dose of 4F-PCC (1,500 or 2,000 units) is just as effective as variable weight-based dosing (25-50 units/kg) at establishing hemostasis for most patients on warfarin or Factor Xa-inhibitors.^[33–37]^

Based on these discussions, recent AHA recommendations, and expert consensus,^[20,21]^ we propose a simplified emergency department ICH algorithm with clear, time-critical treatment targets for anticoagulation reversal, blood pressure control, and emergent neurosurgical interventions (Figure 2). In this proposed ICH treatment paradigm, Code Stroke activation is recommended for all patients with suspected ICH, including those presenting with mild or atypical stroke symptoms, sudden onset of severe headache, ground-level fall, or unexplained unresponsiveness. A non-contrast head CT should be performed within 15 minutes of ED arrival. Once ICH is confirmed on CT, patients with anticoagulation-associated ICH should receive anticoagulation reversal, systolic blood pressure (SBP) reduction to 130-150 mmHg, and neurosurgery consultation within 60 minutes, as outlined in Figure 2. A fixed dose of 4F-PCC (2,000 units) alone for Factor Xa inhibitor-associated ICH or alongside 10 mg of IV vitamin K for warfarin-associated ICH, should be administered immediately after CT confirmation without waiting for laboratory INR or other test results. Additional doses of 4F-PCC or vitamin K may be administered as clinically indicated in patients with supratherapeutic INRs or ongoing bleeding.

**Fig 2.**
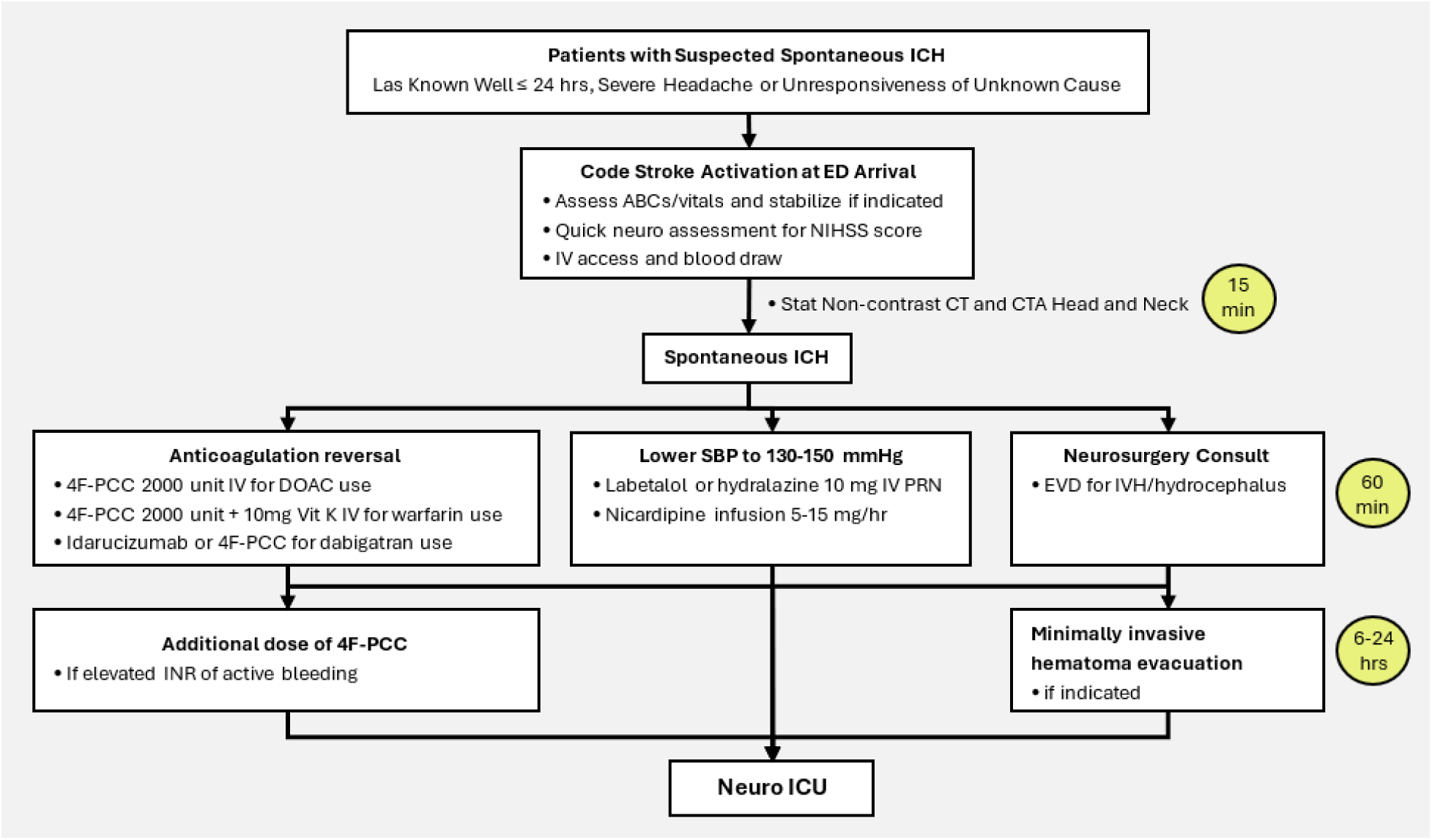
Simplified ED ICH Algorithm

This study has several limitations. First, given the small sample size, our single-center findings may not be generalizable to the broader population of patients with anticoagulation-associated ICH. Second, as expected for a single-center Brief Communication, there is an inherent risk of Type I or Type II errors in the interpretation of baseline demographics and clinical results. For example, the age difference observed between the Code Stroke activation and non-activation groups may represent a false positive (Type I error) due to the limited sample size. Third, due to sample size constraints, multivariable modeling could not be performed; potential confounders (e.g., age, baseline NIHSS, or baseline ICH score) likely influenced the clinical decision to activate Code Stroke. Lastly, despite rigorous screening and chart reviews, the retrospective design and relatively small sample size may have introduced selection bias.

## CONCLUSIONS

Our findings and published evidence support Code Stroke activation for all suspected cases of ICH, the implementation of a time-critical treatment protocol targeting a DTT time of less than 60 minutes, and immediate anticoagulation reversal with fixed-dose 4F-PCC without waiting for laboratory INR results. These represent key operational strategies to optimize the acute management of anticoagulation-associated ICH.

## Author contributions

HT contributed to chart review, data acquisition and manuscript revision. MC contributed to data acquisition, chart review and preliminary statistical analysis. DD contributed to data acquisition and screening anticoagulation-related ICH. JC and MS contributed to data interpretation and manuscript revision. WY contributed to study design, data verification and interpretation, statistical analysis, and drafting and finalizing the manuscript.

## Data Availability

De-identified data are available from the corresponding author upon reasonable request.

